# Spatial patterns of dental disease in patients with low salivary flow

**DOI:** 10.1101/2021.10.04.21264534

**Authors:** Diana M. Proctor, Christof Seiler, Adam R. Burns, Samuel Walker, Tina Jung, Jonathan Weng, Shanne Sastiel, Yoga Rajendran, Yvonne Kapila, Meredith E. Millman, Gary C. Armitage, Peter M. Loomer, Susan P. Holmes, Mark I. Ryder, David A. Relman

## Abstract

Low salivary flow, or hyposalivation, is associated with an increased incidence of dental caries and a shift in their location from biting surfaces towards coronal and root surfaces. However, the relationship between salivary flow and periodontal disease is less clear. To identify clinical indicators of low salivary flow -- including the spatial pattern of dental and periodontal disease, features of the supra- and subgingival microbiota, and symptoms of dry mouth -- we enrolled individuals into two cohorts. The low flow cohort (N = 32) consisted of individuals with a presumptive diagnosis of the autoimmune disorder Sjögren’s Syndrome (SS) while the control cohort (N = 119) consisted of healthy controls. We constructed a series of tooth-specific linear models to quantify the extent to which patient cohort, age, and unstimulated whole salivary flow rate (UWS-FR), independent of each other, are associated with dental and periodontal disease at each tooth. While age and a diagnosis of SS correlated with the site-specific increment of disease so too did UWS-FR. Not only were lower UWS-FRs associated with a greater number of decayed, missing, or filled surfaces at 21 teeth, but they were also associated with increased recession, as measured by clinical attachment loss (CAL), at 10 teeth (adjusted p < 0.05). In addition, we examined microbiota community structure at different tooth sites using data from 427 subgingival and supragingival samples of 6 individuals and found that microbial dispersal is reduced in patients with low salivary flow, but only at supragingival and not at subgingival sites. Finally, we found that complaints by subjects of a negative impact on overall quality of life were associated with a UWS-FR less than 0.1 mL/min. Overall, our results suggest that novel predictors of hyposalivation can be identified by integrating clinical, microbial, and patient history data.

## Introduction

On average, the unstimulated whole salivary flow rate (UWS-FR) of healthy adults ranges between 0.3 and 0.4 mL/min (Becks and Wainwright 1943). Clinically, hyposalivation, or low salivary flow, is defined as an UWS-FR < 0.1 mL/min. Medication is the most common cause of hyposalivation and Sjögren’s Syndrome (SS), a prevalent chronic autoimmune disorder, is a second major cause (von Bultzingslowen et al. 2007). In contrast to patients who experience low salivary flow due to irradiation of cancers involving the head and neck, these two patient populations experience an insidious onset of hyposalivation with delayed diagnoses. Most Sjogren’s patients are diagnosed 6.5 or more years after the onset of xerostomia and nine years after tooth loss has occurred (Baudet-Pommel et al. 1994; Mignogna et al. 2005; Shiboski et al. 2012).

Delays in diagnosing hyposalivation may be due to several reasons. First, the salivary flow rate threshold of 0.1 ml/min is imperfectly correlated with clinical phenotypes. Individuals with flow rates < 0.1 mL/min may not experience signs or symptoms of low salivary flow, while individuals with flow rates as high as 0.3 ml/min may complain of dry mouth (Ben-Aryeh et al. 1985; Dawes 2004). Second, hyposalivation may antedate xerostomia, and patients often suffer from reduced salivary flow for years before it is perceived (Mathews et al. 2008; von Bultzingslowen et al. 2007). Finally, even when patients do report xerostomia to physicians, UWS-FR is not measured in the primary care setting. Collectively these factors lead to diagnostic delays, preventable tooth loss, and an increased burden of caries in medicated patients and in patients with SS (Baudet-Pommel et al. 1994).

Individuals with hyposalivation not only experience more caries, but they also experience a shift in the spatial pattern of dental disease. In the elderly, the risk of root surface caries in the lower jaw increases with the number of prescribed medications with xerostomic effects (Kitamura et al. 1986). Likewise, surgical removal of the salivary glands in rodents leads to an increase in root surface caries affecting mandibular surfaces (Bowen et al. 1988). Compared with healthy individuals, patients with SS are at an increased risk of developing not only root surface, but also coronal caries (Ravald and List 1998). Irradiation of salivary glands in the course of cancer therapy leads to caries on the incisal edges of anterior teeth, cusp tips of posterior teeth, and lingual surfaces (Dreizen et al. 1977). Shifts in the site-specific occurrence of disease in 3 distinct patient populations suggests salivary flow normally functions to protect mandibular root and coronal sites from caries. Moreover, these population level data imply that salivary flow normally plays a role – either directly or indirectly -- in maintaining a beneficial microbiota at these sites since the metabolic activity of the microbial consortia at the tooth surface must change in order for cavitation to occur. When it occurs chronically, the loss of salivary flow leads to a persistent shift in the spatial patterning of the oral microbiota (Proctor et al. 2018).

Uncertainties surrounding the flow rate threshold predictive of increased disease risk highlight the need for new diagnostic indicators, which may be used to predict low flow, before caries occur. Towards that end, we sought to identify features of xerostomia and the microbiome that correlate with flow rate and which may predict hyposalivation, since such features would permit early intervention. Our results suggest that chronic low salivary flow is associated with an increase in DMFS and CAL at a wider array of tooth sites than previously appreciated. In addition, we identify features of the microbiota, including reduced rates of microbial dispersal, and symptoms of dry mouth that may serve as indicators of hyposalivation. These observations are significant since they indicate new avenues for diagnosing hyposalivation, a condition for which UWS-FR is imperfectly sensitive and specific.

## Results

### Clinical and demographic features of the patient population

The complete set of inclusion and exclusion criteria for each cohort is provided as supplementary data **(Supplementary Methods)**. Data were collected from 151 participants in 2 cohorts: the low flow cohort (N = 32) consisted of individuals with a presumptive diagnosis of SS while the control cohort (N = 119) consisted of healthy controls. Gender identity, race, and ethnicity did not significantly differ between the two groups (Chi-square tests, p > 0.1) **(Table S1)**. The majority of subjects in both cohorts were White or Asian and did not identify as Hispanic or Latino. The proportion of females in the low flow group did not significantly differ from that of the controls (Chi-square test, p = 0.2), though substantially more females than males were enrolled in both cohorts. Roughly 93% of the low flow cohort identified as female, consistent with the known sex bias in the occurrence of SS. Likewise, SS tends to be diagnosed in middle to late life, and the average age of individuals in the low flow cohort (mean, 60.8 years) was higher than the average age of controls (mean, 32.6 years). Despite a mean difference of 28 years, there was considerable overlap between the two groups (**Figure 1a**; 95% confidence intervals (CIs) surrounding the unpaired mean difference, 23.8-32 years). Indeed, the controls appeared to segregate into 2 groups, the largest group consisting of subjects under the age of 40 and the second group reflecting our push to enroll controls similar in age to the SS patients. Despite our best efforts, just 20% of the controls sampled in this study were over the age of 40.

**Figure 1.**
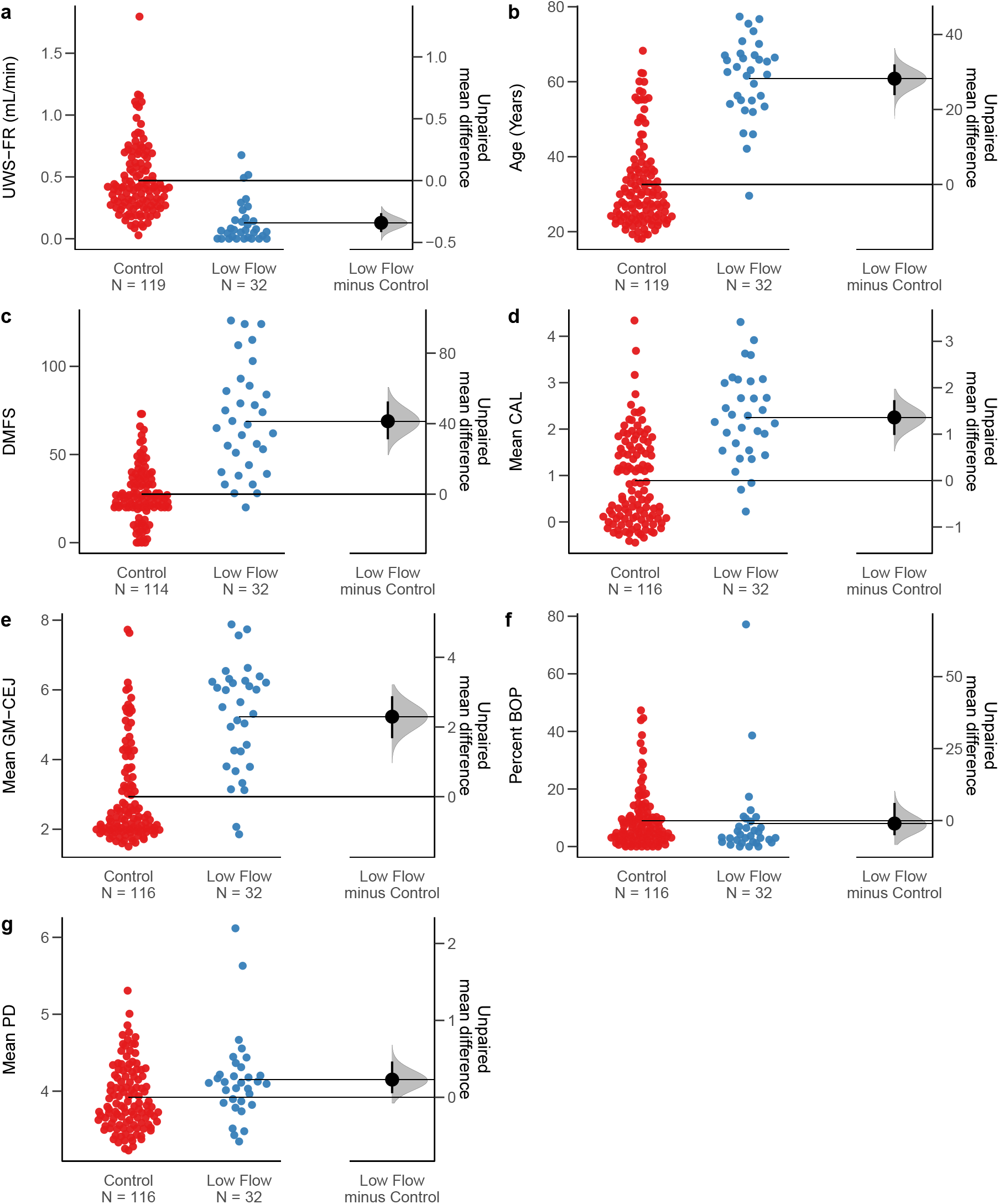
Gardner-Altman estimation plots reveal significant demographic and clinical differences between low flow and control cohorts. Data for control and low flow cohort patient Swarm plots of a) age, b) UWS-FR, c) DMFS, d) CAL, e) GM-CEJ, f) BOP, and g) PD are plotted on the left-aligned axis. The right-aligned axis displays the point estimate of the unpaired mean difference between groups, surrounded by their 95% confidence intervals (95% CI).

As expected, based on inclusion and exclusion criteria, UWS-FRs varied significantly between the two groups **(Figure 1b)**. The mean UWS-FR of patients in the control cohort (0.470 mL/min) was on average 0.335 mL/min higher than the mean rate of the low flow cohort (0.129 mL/min) (95% CI, 0.253--0.410 mL/min). Given that we excluded individuals with active disease, all measures of disease reflect a past history of dental disease.

Further, the incidence of past dental disease was higher in the low flow cohort compared to the controls. The mean difference in decayed missing filled surfaces (DMFS) between cohorts differed significantly from 0 (**Figure 1c**) (95% CI, 30.4-52.4) with a mean DMFS of 27.5 in controls and mean DMFS of 68.9 in the low flow cohort. Similarly, mean clinical attachment loss (CAL) was significantly higher in the low flow cohort (2.25) compared to the controls (0.891) with a mean difference of 1.38 (**Figure 1d)** (95% CI, 1.01-1.74). The gingival margin cemento-enamel junction (GM-CEJ) was also higher in the low flow cohort (mean, 5.23) compared to controls (mean, 2.93) with a substantial mean difference of 2.3 (**Figure 1e**) (95% CI, 1.68-2.88). Despite higher measures of DMFS, CAL, GM-CEJ in the low flow cohort, bleeding on probing (BOP) and probing depth (PD) did not significantly differ between the two groups suggesting differences in GM-CEJ and CAL reflected past or chronic periodontal disease rather than active disease **(Figures 1f, 1g)**.

### Age and cohort exert independent effects on the spatial pattern of dental disease

Given that age is known to be correlated with the prevalence of dental disease (Algarni et al. 2018; Kassebaum et al. 2017) we sought to evaluate the relationship between salivary flow and site-specific prevalence of dental disease while controlling for age. We used regression coefficients to quantify the extent to which the site-specific increment of disease (DMFS, CAL, GM-CEJ, BOP, PD) varied depending on a one-unit change in each of several standardized predictors (age, UWS-FR, cohort, and the interaction between cohort and UWS-FR) while holding the other 3 predictors constant.

A site specific DMFS increment was directly correlated with age across virtually all teeth (adjusted p < 0.05) excluding 3 anterior teeth (teeth 6, 24, 27). The site-specific effects of age can be discerned by comparing the regression coefficients for each tooth to each other. At tooth 8, DMFS increased by 0.076 Ordered Quantile (ORQ) transformed surfaces for each year of life while at tooth 15 DMFS increased by 0.289 QRQ surfaces, a rate approximately 3.8 times higher **(Figure 2a)**. Indeed, DMFS coefficients for age were about 3-20 times higher at posterior compared to anterior sites, consistent with a higher rate of attack for posterior compared to anterior teeth **(Figure 2a)**. Similarly, age was a significant and direct predictor of CAL **(Figure 2b)**, and GM-CEJ **(Figure 2c)** for all tooth sites (adjusted p < 0.05). While coefficients for periodontal measures tended to be higher at posterior versus anterior teeth, in both jaws, coefficients did not clearly segregate into anterior and posterior groups, as they did for DMFS. In contrast to CAL and GM-CEJ, BOP and PD were correlated with age at a limited number of tooth sites. BOP **(Figure 2d)** was correlated with age at 4 mandibular sites (teeth 19, 23, 24, 30) and 1 maxillary site (tooth 2) while PD **(Figure 2e)** was significantly correlated with age at 3 maxillary sites (teeth 2, 5, 15) and 1 mandibular site (tooth 24).

**Figure 2.**
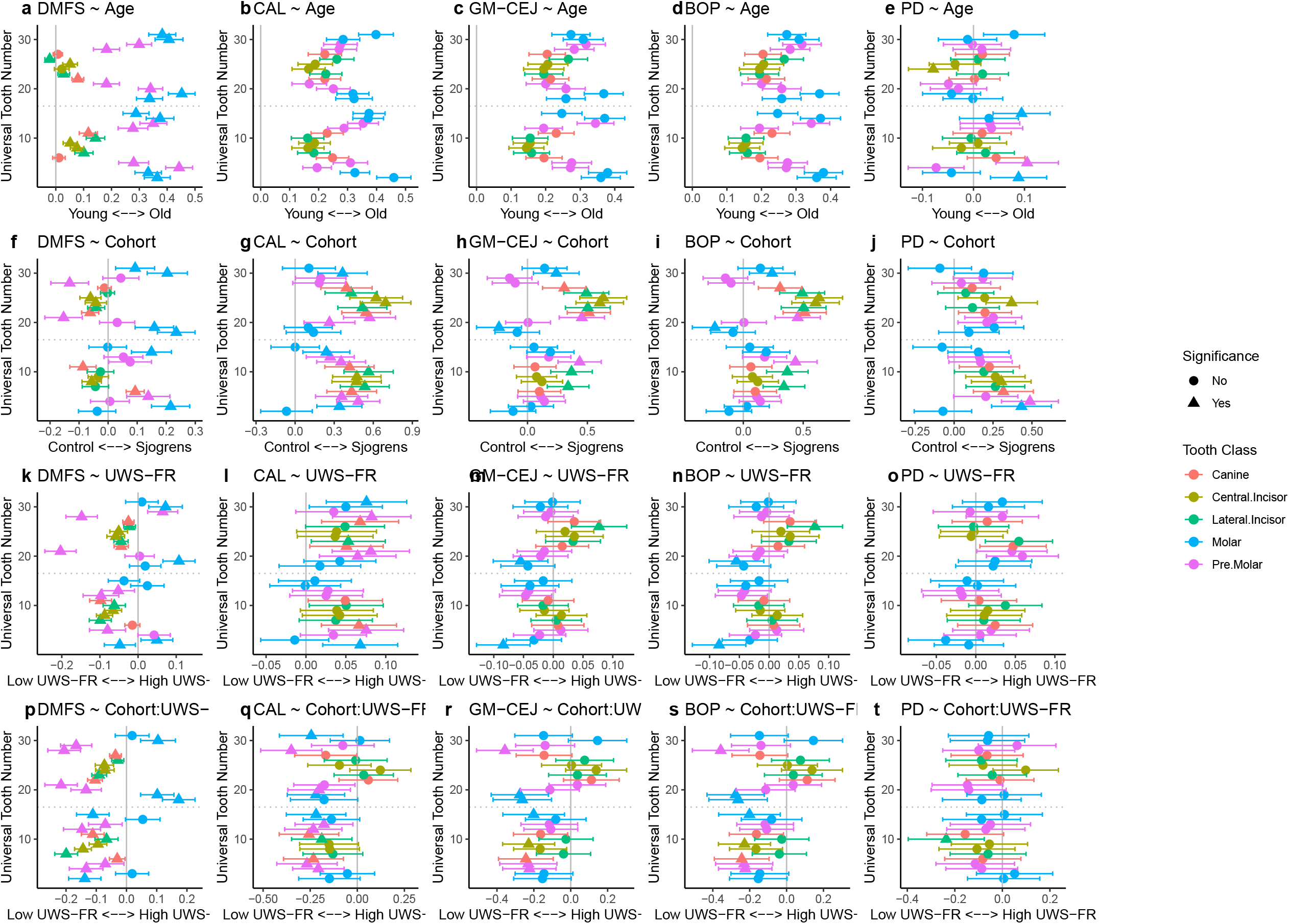
Spatial modeling reveals independent effects of age and patient cohort on the spatial pattern of dental disease. Coefficients with 95% confidence intervals are plotted as a function of universal tooth number for a series of per-tooth linear models where DMFS (a, f, k, p), CAL (b, g, l, q), GM-CEJ (c, h, m, r), BOP (d, i, n, s), and PD (e, j, o, t) were regressed against age, patient cohort, UWS-FR and the interaction between UWS-FR and cohort. Intercepts were calculated, but not visualized for each model. Colors map to different tooth classes. Triangles denote estimates with adjusted p-values < 0.05, while squares denote estimates with adjusted p-values > 0.05. P-values were adjusted by controlling the false discovery rate (FDR) at 5% using the Benjamini-Hochberg (BH).

Next, we sought to examine the independent contribution of cohort, or having a presumptive diagnosis of SS, on the site-specific increment of dental disease. Across all disease metrics, the regression coefficients for cohort were smaller than those for age, suggesting that cohort has a smaller effect size than age, though it still explains variation in the occurrence of dental disease. With a categorical variable as the predictor, the regression coefficients represent the difference in the average disease increment between the control and low flow cohorts. In the low flow cohort, 16 teeth had significant coefficients that could be distinguished from zero **(Figure 2f)**. Negative coefficients were observed at several anterior teeth in the maxilla (teeth 6, 8, 11) and mandible (teeth 22, 23, 24, 25). In addition, while both groups tended to have more caries at posterior sites, low flow subjects had between 0.09 and 0.23 more QRQ DMFS per tooth at maxillary teeth 3, 5, 6, and 14, and mandibular teeth 18, 19, 30, and 31, as compared to the reference group.

After adjusting for multiple testing, CAL differed significantly at 21 sites between patient cohorts **(Figure 2g)**. Compared to controls, recession was on average 0.48 QRQ mm (range, 0.41-0.56 QRQ mm) greater in the low flow cohort at anterior maxillary teeth (teeth 6, 7, 8, 9, 10, 11) compared to controls, again holding all other predictors constant. Similarly, CAL was greater by an average of 0.53 QRQ mm at anterior mandibular (teeth 22, 23, 24, 25, 26, 27) sites, 0.34 mm at posterior maxillary (teeth 3, 4, 5, 12, 13, 14) and 0.39 QRQ mm at posterior mandibular (teeth 20, 21, 30) sites. In contrast to CAL, GM-CEJ tended to be greater in the low flow cohort compared to controls at a more limited number of teeth **(Figure 2h)**, including just 9 teeth in the mandible (teeth 19, 21, 22, 23, 24, 25, 26, 27, 30) and 3 in the maxilla (teeth 7, 10, 12). BOP was not significantly correlated with cohort at any site **(Figure 2i)** while patients with low flow had higher PD at predominantly maxillary sites (teeth 3, 4, 6, 8) though one mandibular site (tooth 24) also reached significance **(Figure 2j)**.

Since we enrolled individuals with a presumptive diagnosis of Sjogren’s Syndrome in our low flow cohort, effects related to cohort could be attributable to a reduction in salivary flow as well as changes in salivary composition or immune function. To assess the explicit effect of flow rate on dental disease, we also included UWS-FR in our models, allowing us to see its contribution independent of either age or cohort. Compared to age and cohort, coefficient sizes for UWS-FR were universally smaller across all disease metrics, indicating an overall smaller effect of flow rate on disease outcomes. Despite a relative difference in the magnitude of effect sizes, UWS-FR was significantly correlated with DMFS, CAL, and GM-CEJ at a variety of different tooth sites. In particular, 21 tooth sites **(Figure 2k)**, including anterior maxillary (teeth 7, 8, 9, 10, 11), posterior maxillary (teeth 2, 3, 5, 12, 13), anterior mandibular (teeth 22, 23, 24, 25, 26, 27), and posterior mandibular (teeth 19, 21, 29, 30) sites. Whether in the maxilla or mandible, anterior sites tended to have negative correlation coefficients indicating that as UWS-FR increased the number of DMFS decreased at these sites. In contrast to the wide variety of sites exhibiting a correlation between DMFS and UWS-FR, CAL was correlated with UWS-FR at a smaller number of sites **(Figure 2l)** including 3 sites in the upper right jaw (teeth 2, 5, 6) and 7 sites in the lower jaw (teeth 20, 21, 22, 23, 27, 27, 31). Even fewer sites exhibited a correlation between GM-CEJ and UWS-FR **(Figure 2m)**, including 2 molars (teeth 2, 19) and 1 incisor (tooth 26). BOP was correlated with UWS-FR at just on tooth site (tooth 9; **(Figure 2n**) while PD was not correlated with UWS-FR at any site **(Figure 2o)**. Taken together, these data suggest that in patients with a presumptive diagnosis of Sjogren Syndrome there is a detectable increase in the site-specific increment of caries and periodontal disease that is attributable to salivary flow rate.

Next, we explored the interaction between cohort and UWS-FR. In particular, we were interested in knowing whether the direction or magnitude of the effect of UWS-FR differed between the reference cohort (i.e, controls) and the low flow cohort. Out of 25 significant coefficients for the interaction between cohort and UWSFR as a predictor of DMFS, 22 were negative. The mandibular molars were the exception (tooth 18, 19, 30) with coefficients that ranged between 0.10-0.17 **(Figure 2p)**. Consistent with this finding, we observed in a less sophisticated model that as UWS-FR decreased in the low flow cohort the DMFS increment increased at a greater rate than the DMFS increment increased with reductions in UWS-FR in the control cohort **(Supplementary Figure 1)**. Several coefficients for the interaction between cohort and CAL were also significant in the maxilla (teeth 4, 5, 6, 10, 11, 12, 13, 15) and mandible (teeth 19, 20, 28, 31) **(Figure 2q)** with similar patterns of disease observed when considering GM-CEJ **(Figure 2r)**. The interaction between cohort and UWS-FR was not significantly correlated with BOP **(Figure 2s)** or PD **(Figure 2t)** at any site.

### Impact of age and low salivary flow on the microbiota

To examine whether the differences in past dental disease were associated with shifts in the oral microbiota we next analyzed the relationship of age, cohort, and UWS-FR to structure of the subgingival microbiota from each of 3 control and 3 low flow patients. Subgingival microbiota analysis from the first molars, canines, and central incisors was integrated with previously-published data from the supragingival microbiota at these same teeth in these same 6 patients (Proctor et al. 2018).

Differences in structure of the supragingival and subgingival communities, based on Bray Curtis dissimilarity, explained segregation of samples along axis 1, which accounted for 17.6% of the variation **(Figure 3)**. UWS-FR explained differences among subgingival and supragingival communities along axis 2, which accounted for 12.8% of the variation in the data **(Figure 3a)**. On the other hand, age co-segregated with UWS-FR at subgingival but not at supragingival sites **(Figure 3b)**. Supragingival sites scored as 1 or 2 DMFS clustered together in the low flow cohort segregating from supragingival sites with 0 DMFS **(Figure 3c)**. At subgingival sites, samples from younger control subjects tended towards positive scores along axis 2 while samples from the older low flow subjects tended towards neutral to negative scores along axis 2. At supragingival sites, the one older control subject’s (55.2 years) supragingival samples clustered with the two young control subject’s communities (23 and 28 years). Similar results were obtained when using different distance metrics **(Supplementary Figure 2)**. Examining axis 1 scores as a function of tooth class revealed that subgingival communities, unlike supragingival communities, do not differ by tooth class in either the low flow or control cohorts **(Supplementary Figure 3)**.

**Figure 3.**
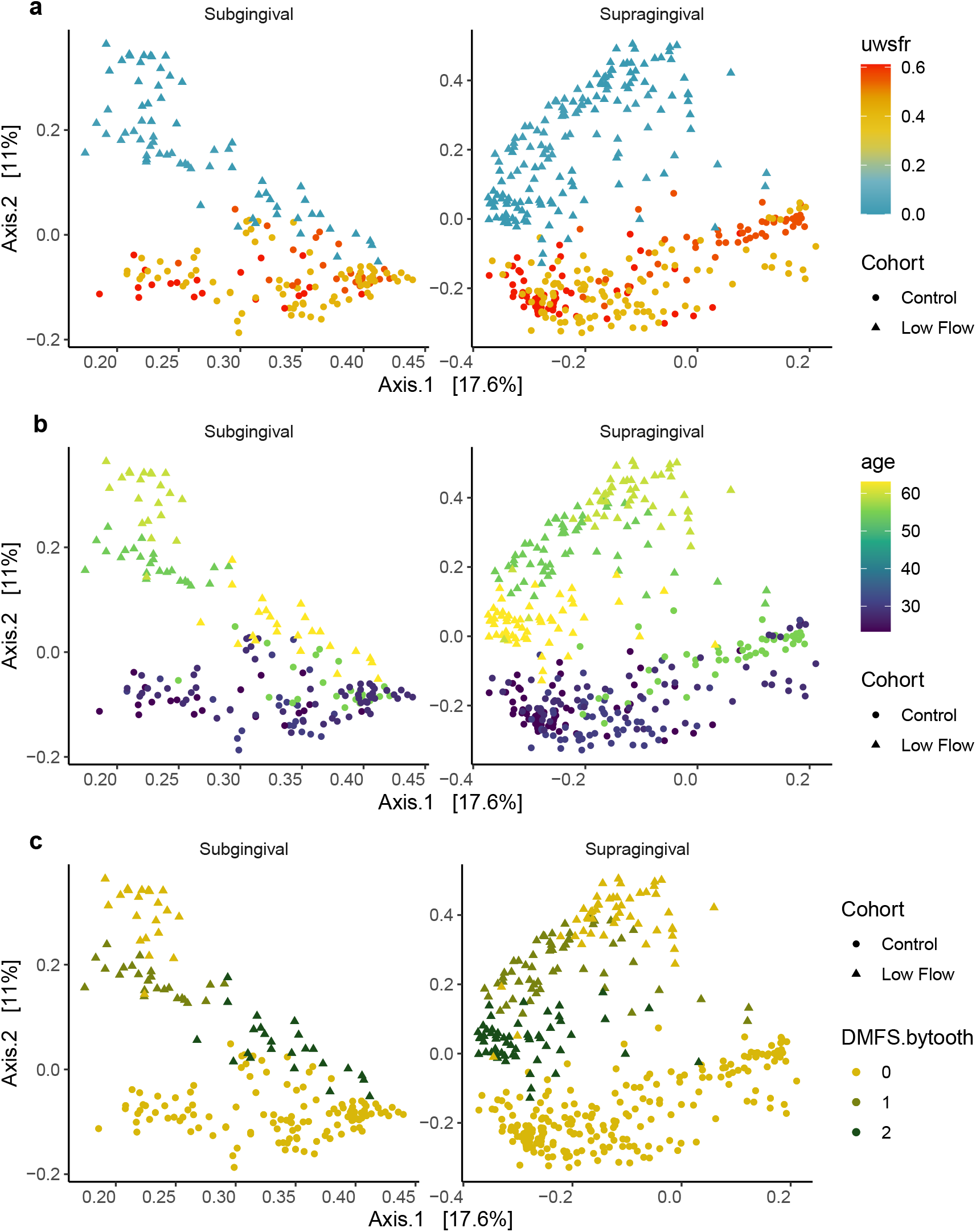
Subgingival and supragingival communities from 3 control and 3 low flow subjects segregate by UWS-FR while age imperfectly separates supragingival communities. Principal coordinates analysis on Bray Curtis dissimilarity was performed on the combined subgingival and supragingival dataset. The PCoA is displayed as a facet wrap with subgingival and supragingival samples in left and right panels, respectively. a) Communities segregated by UWS-FR along the second coordinate which explained 12.8% of the variance. b) Samples clustered by age for subgingival, but not supragingival sites. c) Supragingival communities from sites within low flow subjects with 1 or 2 DMFS segregated from sites with 0 DMFS.

Constrained correspondence analysis revealed that age, GM-CEJ, PD and CAL were correlated with each other and explained the segregation of low flow samples with positive scores along axis 1, consistent with the site-specific regression models **(Figure 2)**. Subgingival and supragingival sites from subject 3-303 had higher CAL and lower DMFS values than sites from subject 3-301. At the same time, subgingival and supragingival sites from subject 3-301 were enriched in *Streptococcus* and *Veillonella* species (ASV12, ASV4, ASV7, and ASV8) and distinct from sites that were enriched in *Prevotella denticola* (ASV11) along axis 2. Samples from control subjects with higher UWS-FR had negative scores on axis 1 along with samples from one low flow subject, 3-302 who had the highest DMFS scores among these subjects **(Supplementary Figure 4)**. Taken together, these pilot data suggest that the composition of the microbiota is correlated with age, cohort, and flow rate, potentially reflecting correlations between the incidence of disease at different sites related to these predictors.

### Low salivary flow is associated with an increase in rates of microbial migration

In our prior work, we observed that communities inhabiting exposed tooth surfaces of different tooth classes could be differentiated from one another in controls, but not low flow subjects. Here, we report that subgingival communities do not differ based on tooth class in either cohort, in these 6 individuals. Based on these observations, we hypothesized that reductions in salivary flow result in a net increase in microbial dispersal, defined here as an increase in the movement or retention of microbes between sites, at supragingival sites. To test this hypothesis, we used two models of microbial dispersal, investigating whether rates of microbial migration differ among tooth habitats (subgingival, supragingival) within an individual as well as between individuals based on cohort (i.e., in 3 low flow patients vs. 3 controls).

Regardless of cohort, migration rates were higher at supragingival compared to subgingival sites **(Figure 4, Table S2**). In addition, migration rates were higher in patients with low salivary flow compared to controls, suggesting that salivary flow moderates the rate of migration in the healthy human oral cavity. Further, we observed a significant interaction between participant cohort and the gingival habitat, with a substantial difference in migration between the two cohorts at supragingival sites with a less pronounced difference at subgingival sites. These findings were robust to the method used to infer migration **(Supplementary Figure 5; Supplementary Results)** and suggest an increased rate of migration across the supragingival area of the mouth in the low flow cohort.

**Figure 4.**
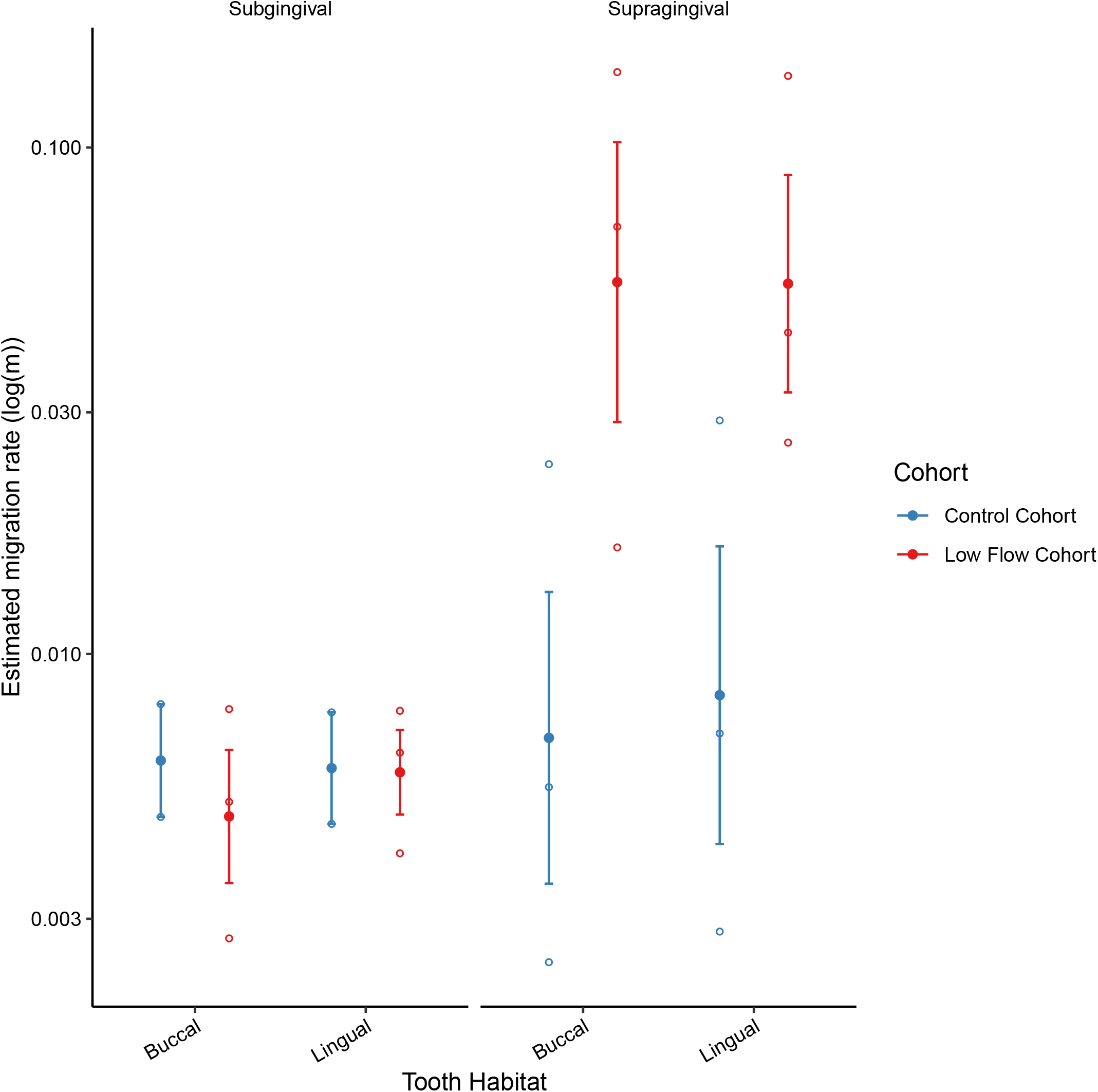
Estimated migration rates by habitat within each individual. Rates were inferred using the Sloan Community Neutral Model. Each hollow point represents the estimated migration rate for each of the defined metacommunities for each individual, while the solid point represents their average with standard error bars. Colors correspond to patient cohort.

### Subjective predictors of low salivary flow

Next, we investigated the extent to which complaints of dry mouth are predictive of low salivary flow using a previously developed Visual Analog Scale (VAS) (Pai et al. 2001) containing six questions on quality of life and feelings of dry mouth, which we adapted by the addition of a question concerning the extent to which patients experienced dryness of the hard palate. Members of the low flow cohort complained of impaired ability to swallow due to dry mouth, impaired overall quality of life, impaired ability to speak, as well as feelings of dryness impacting the throat, tongue, lips and hard palate (p < .01, **Supplementary Figure 6**).

Individuals rated their responses to questions, on the VAS, based on the perceived severity of their symptoms. A principal component analysis of these ratings was performed to identify questions whose responses could segregate patients based on the severity of their complaints of dry mouth **(Figure 5a)**. The first axis explained 86% of the variation in the data and principally segregated patients into the low and “not low” cohorts with low flow subjects sharing positive scores along axis 1. The 12 control subjects who grouped with the low flow screening cohort also tended to have a lower UWS-FR than the 61 control subjects who clustered together, though the difference in means was not significant. A conditional inference tree was used to identify the symptomatic complaints that were most effective at separating patients into low flow and not low flow cohorts **(Figure 5b)**. While these results should be validated on a larger dataset, 84.6% of subjects (11/13) in this study who indicated that low salivary flow negatively impacted their overall quality of life (VAS ≥ 56) had flow rates of less than 0.1 ml/min. On the other hand, most subjects who rated a negligible impact of low flow on their quality of life as well as an absence of dryness on their lips had flow rates exceeding 0.1 ml/min. The AUC of our model exceeded 84% with 10-fold cross validation. These same significant predictors were also identified using the random forest algorithm as the most discriminative of patients in the low flow versus the control cohort **(Supplementary Figure 7)**.

**Figure 5.**
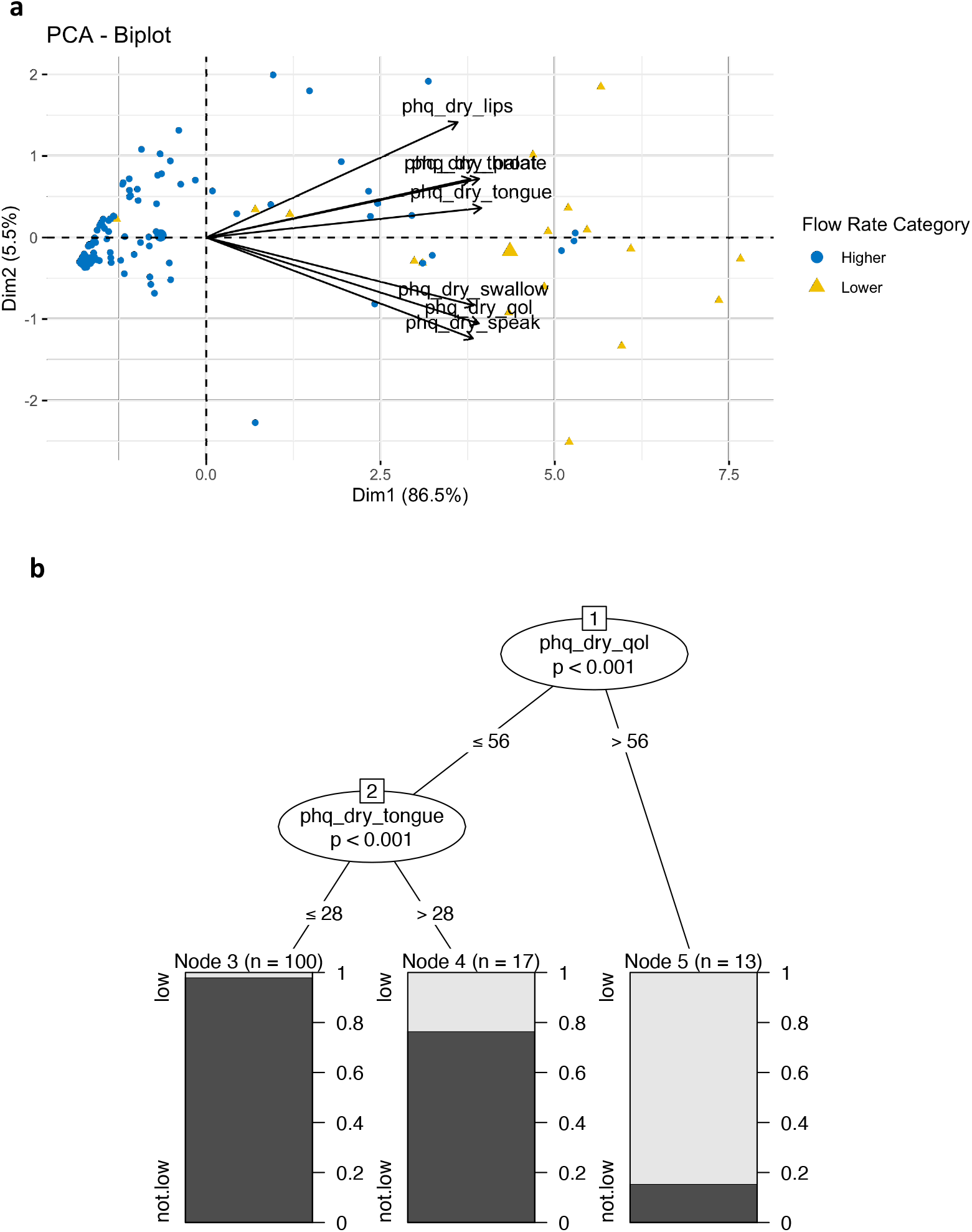
Symptoms of dry mouth predict low salivary flow. a) Principal component analysis of subject responses to visual analog scale revealed two groups of questions – questions that segregate subjects who complain of impacts to quality of life (dry_speak, dry_qol, dry_swallow) and questions that indicate feelings of dry mouth (dry_tongue, dry_throat, dry_palate, dry_lips). b) Conditional inference tree was used to identify the variables most predictive of separating patients into groups based on low (UWS-FR < 0.1ml/min) and not low (UWS-FR > 0.1 ml/min) subgroups.

## Discussion

Hyposalivation is associated with reduced quality of life, including reduced oral health quality of life, even in patients whose dental health is well managed subsequent to diagnosis. Given that patients with hyposalivation are typically not diagnosed until they have lost a first tooth to disease, a critical need in this patient population is the development of methods to identify low salivary flow before the onset of dental caries, so that interventions can be implemented to prevent deterioration of oral health quality of life. Our goal was to identify microbial and symptomatic predictors of salivary flow, analyzing multi-facetted data on the same patient population.

Aging is associated with spatial patterns in the incidence of dental disease, impacting surfaces above and below the gumline (Algarni et al. 2018; Kassebaum et al. 2017). Gingival recession increases as a function of age, exposing root surfaces which become vulnerable to caries. UWS-FR has been shown to decrease as a function of age in individuals taking medications as well as in the otherwise healthy elderly (Fure and Zickert 1990). Prior work suggests that the function of the submandibular glands decreases with age, with reductions in UWS and SWS secretions averaging between 22% to 39%, respectively (Baum 1981; Pedersen et al. 1985). Reductions in salivary flow may explain the increase in the incidence of root surface caries subsequent to the sixth decade of life (Sumney et al. 1973). Our data support the hypothesis that aging and the effects of chronic low salivary flow exert cumulative effects on the shifting spatial pattern of dental disease in patients with hyposalivation.

Dental diseases can be considered ecological catastrophes that leave longstanding impressions on the oral ecosystem (Marsh 2006). Since restoration of dental caries fails to eliminate cariogenic bacteria from smooth surface margins, restored surfaces may serve as reservoirs for reinfection of sites adjacent to or distant from the restoration (Featherstone 2000). Current evidence indicates patients with low salivary flow have a microbiota enriched in cariogenic bacteria (Almstah et al. 2003; Almstahl et al. 2001; Eliasson et al. 2006; Proctor et al. 2018). Here, we report that patients with hyposalivation experience a higher number of decayed, missing and filled surfaces, and thus harbor more reservoirs for cariogenic bacteria, and that they also experience an increase in detectable rates of microbial migration.

We hypothesize that the increased rate of migration at supragingival surfaces may underlie the increased risk of dental caries patients with hyposalivation experience post-restoration and despite fastidious compliance with dental care regimens (Segal et al. 2009). Consistent with *in vitro* studies of shear force (Fernandez et al. 2017), we propose that our preliminary data suggest shear associated with salivary flow may control the diversity and composition of the supragingival biofilm *in vivo* by limiting dispersal. Under normal circumstances, microbes which detach from supragingival surfaces are marshalled out of the oral cavity through the combined effects of abrasion from the tongue, salivary flow, and deglutition. We propose that following detachment, where there is chronic suppression of flow rates, microbial species are able to successfully attach and grow as part of the tooth-associated supragingival biofilm at sites distal to their origin. Thus, salivary flow may hinder the process of attachment and growth at supragingival sites. In contrast to supragingival sites, our preliminary data suggest that salivary flow does not exert top-down control over the rate of dispersal between subgingival sites.

Dispersal between subgingival sites may be controlled more by the rate of exudation of gingival crevicular fluid from the gingival sulcus to the gingival margin than by the rate of salivary flow. Moreover, desquamation of the epithelium may also limit dispersal into the subgingival crevice. Alternatively, out of 12 teeth that we sampled only 4 teeth (tooth 8 and 9 or 24 and 25) had sites immediately adjacent to each other. As a result, if dispersal between subgingival sites occurs at a greater frequency between adjacent teeth than between distal teeth our power to detect differences in rates of migration between subgingival sites in controls vs. low flow subjects may have been limited, particularly when considering our overall small sample size. Future studies that survey a larger number of teeth at sites above and below the gumline are needed to validate these observations.

Diagnostics that identify patients who may be suffering from occult low salivary flow should mitigate risk of recurrent dental disease in these patients. We identified two symptomatic complaints as the most discriminative, identifying salivary flow rates < 0.1 mL/min with ∼85% accuracy. These symptomatic complaints – the extent to which dry mouth negatively impacted overall quality of life, and the extent to which dryness was felt on the tongue – were as effective at distinguishing different microbial communities as UWS-FR, suggesting that, in our cohort, flow rate and xerostomic complaints are highly correlated. Our findings are consistent with prior work suggesting that functional impairment is the most discriminative xerostomic complaint for separating patients based on salivary hypofunction (Sreebny and Valdini 1988). Age related xerostomic complaints are known to markedly increase in the 6^th^ decade of life with women more frequently reporting symptoms of dryness compared to men (Niklander et al. 2017). In response to anticholinergic suppression of salivary flow, older adults experience more prolonged functional impairment, as measured by both flow rate and symptomatic complaints of xerostomia, with a longer duration of difficulties speaking and swallowing due to dryness, as well as longer periods of dryness reported on the lips. Future large-scale prospective studies should endeavor to examine the relative onset of xerostomic complaints and hyposalivation over the decades of human life.

Our study was limited by a design that enrolled only patients having low salivary flow due to a presumptive diagnosis of SS, rather than including patients who experienced low salivary flow due to a wide variety of conditions. This study design resulted in an observational study with a relatively small sample size and an uneven distribution of ages with a younger control cohort and an older low flow cohort. While these challenges can be addressed statistically an ideal design would include age matching between participant cohorts. Despite these limitations our work provides a framework, including a script for reproducibility, that can be used by others to integrate the analysis of microbiome data with clinical data and patient histories.

## Methods

### Human Subjects

Subjects were enrolled into a larger study of hyposalivation. Written, informed consent was obtained from all participants prior to dental examination or sample collection in compliance with human subjects protocols approved by the University of California, San Francisco (UCSF) Human Research Protection Program and Institutional Review Board (Protocol 11-06273), and the Stanford University Administrative Panels on Human Subjects in Medical Research (Protocol 21586). Subjects were recruited into 2 cohorts: (1) 152 healthy adults were recruited into a “control cohort”; and (2) 32 individuals who experienced low salivary flow due to the autoimmune disorder, SS, were recruited into a “low-flow cohort”. A complete description of clinical data measurements and sample processing workflows are included in **Supplementary Methods**.

### Statistical analysis of clinical and demographic data

The `dabestr` package in R (Ho et al. 2019) was used to assess between-group differences in clinical and demographic features. A bootstrap confidence interval was constructed surrounding estimates of the mean difference between the groups. Bootstrap resampling was used to compute assumption-free confidence intervals. Bias correction and acceleration were used in cases of skewness. Between-group differences in ethnicity, race and gender were evaluated using chi-square tests.

### Evaluation of hyposalivation effects on spatial distribution of oral disease

A series of site-specific generalized linear models were evaluated to determine the spatial distribution of oral disease (DMFS, CAL, PD, GM-CEJ, and BOP) with respect to the following predictors: age, cohort, UWS-FR, and cohort:UWS-FR (interaction between cohort and UWS-FR). Response variables were transformed using the Ordered Quantile (ORQ) normalization transformation with the `orderNorm` function within the package `bestNormalize`. This model was applied to each tooth (excluding third molars) using the whole tooth average measurement of each clinical variable. The confidence intervals of each model were plotted to assess the extent to which each predictor explains the spatial patterning of dental disease. P-values, assessing whether coefficients could be distinguished from 0, were adjusted by controlling the false discovery rate (FDR) at 5% using the Benjamini-Hochberg (BH) (Benjamini and Hochberg 1995).

### Analysis of oral microbial communities

Supragingival samples from 3 low flow and 3 control patients previously described (SRA SUB10454805) were analyzed with newly generated subgingival data from the same patients, yielding a total of 427 samples. A complete description of statistical approaches to the analysis of the microbiome is included in **Supplementary Methods**.

### Analysis of an adapted visual analog scale (VAS)

Subjects were asked to use a visual analog scale to rate on a scale of 0 to 100 the extent to which they felt dry mouth negatively impacted their ability to swallow, their overall quality of life, and their ability to speak. In addition, subjects were asked to rate on a scale of 0 to 100 the overall dryness of their throat, tongue, lips, and hard palate. The specific questionnaire is provided as Supplementary Data. A total of 106 individuals responded to the survey, including 73 controls and 24 low flow individuals. Responses from all individuals were analyzed by principal components analysis using the `prcomp` function in base R. A classification tree was generated using the `rpart` function in the `rpart` package.

## Data Availability

The data supporting the results of this study are available in the NIH Short Read Archive, under SRA accession number SRP126946 (http://www.ncbi.nlm.nih.gov/sra/22016164-22016299). The code and data that were used to generate these findings can also be found at https://github.com/dmap02/spatial-pattern-dental-disease. All other data supporting the findings of this study are available within the article and its Supplementary Information files, or are available from the authors upon request.

## Acknowledgements

We thank colleagues at Stanford and at UCSF who provided technical support for this project, including Swetha Kanukula, Lakshmi Karayil, Muneet Shoker, Divya Vadlamudi, Saba Dolatshahi, Anchita Venkatesh, Nicole Davis, and Felix Chen. This work was supported by the National Institutes of Health (R01DE023113 to D.A.R.), the Chan Zuckerberg Biohub Microbiome Initiative (D.A.R.), and by the Thomas C. and Joan M. Merigan Endowment at Stanford University (D.A.R.). Survey data were collected and managed using REDCap electronic data capture tools hosted at Stanford University.

## Author contributions

DMP, PMP, GCA, MIR, DAR, SPH: Conceived of study/developed IRB protocols. DMP: Wrote the first draft of the manuscript. MEM: performed literature review. DMP, CS, ARB, SW, TJ, JW, SS, MM: performed data analysis. YR, MIR, DAR, SPH: assisted with the interpretation of data. CS and SPH guided statistical analysis. All authors reviewed and approved of the manuscript.

