## Supplementary Methods, Tables, Figures for "Spatial patterns of dental disease in patients with low salivary flow"

### Supplementary Data

#### Supplementary data file 1

Supplementary data file 1 is a .xlsx file that can be used to build a duplicate version of the Visual Analog Scale deployed in this study in REDCap.

#### Supplementary data file 2

Supplementary data file 2 is an Excel workbook that contains 2 integrated datasets.

- ASV table, taxonomy file, and sample data mapping file for supragingival samples described in Proctor et al (2018) and the subgingival samples, generated for this manuscript.

#### Supplementary data file 3

Supplementary data file 3 contains the code and figures used to analyze the dataset.

### Supplementary Methods

#### Inclusion and exclusion criteria

Sjogren's Syndrome (SS) subjects were included in the study if they were adults over 18 years old, complained of dry mouth, and had been diagnosed at least three months ago with Sjogren's Syndrome. The diagnosis date of SS was also collected verbally and verified with subjects' documentation. Otherwise, healthy control subjects were included in the study if they were healthy, non-smoking adults over the age of 18 years.

Exclusion criteria for the SS group were 1) having fewer than 15 non-implant teeth; 2) smoke or use chewing tobacco or snuff or quit using tobacco products within the 6 months preceding enrollment; 3) being treated by a physician for an uncontrolled chronic medical condition; 4) have symptoms of or treatment of asthma or acid reflux in the last 3 months; 5) history of radiation therapy to the head or neck; 6) history of oral, systemic antibiotics or antifungals use within the 6 month period preceding enrollment; 7) required to take antibiotics before dental treatment; 8) history of stimulant or heroin abuse or of eating disorders; 9) lactating, pregnant, or intending to become pregnant; 10) any dental treatment during the one month period preceding enrollment and cannot or will not abstain from dental treatments during their enrollment; 11) have fixed dental appliance (retainers, fixed dentures, braces, orthodontic wires); 12) periodontitis, candidiasis, halitosis, tooth pain, or any other disease in the mouth (to patient's knowledge); 13) diagnosed with Sjogren's syndrome by the American European Consensus Criteria or the American College of Rheumatology Classification Criteria fewer than 3 months prior to the date of enrollment.

Exclusion criteria for the control group were 1) having fewer than 15 natural, non-implant teeth; 2) missing both central incisors, both canines, or both first molars in either the maxilla or the mandible; 3) crown or implant replacing both central incisors, both canines, or both first molars in either the maxilla or the mandible; 4) currently being treated by a physician for any chronic medical condition, including asthma and acid reflux; 5) history of radiation therapy to the head or neck; 6) take any medication on a daily basis other than birth control; 7) history of oral, systemic antibiotics or antifungals use within the 6 month period preceding enrolment; 8) required to take antibiotics before dental treatment; 9) history of stimulant or heroin abuse or of eating disorders; 10) lactating, pregnant, or intending to become pregnant; 11) any dental treatment during the 1 month period preceding enrollment and cannot or will not abstain from dental treatments during their enrollment; 12) fixed dental appliance (retainers, fixed dentures, braces, orthodontic wires); 13) experienced dry mouth for a full week at any time in the past 6 months; 14) periodontitis, candidiasis, halitosis, tooth pain, or any other disease in the mouth (to patient's knowledge).

#### Clinical Data Measurements

A calibrated dentist performed a comprehensive dental exam to evaluate the oral and dental health status of each subject. Dental health was evaluated by measuring decayed, missing, and filled surfaces (DMFS), probing depth (PD), gum recession (GM-CEJ), and bleeding on probing (BOP). These measurements were taken at the mesiobuccal, midbuccal, distobuccal, mesiolingual, midlingual, and distolingual sites on each tooth (except third molars) using a North Carolina probe. Clinical Attachment Loss (CAL) was calculated using the following formula: PD + GM-CEJ. Mucosal surfaces in the oral cavity were examined for any sign of disease (e.g., oral candidiasis). No subject had active dental disease at the time of sample collection.  No subject had used antibiotics in the 6 months preceding enrollment.

Unstimulated whole salivary flow rate (UWS-FR; mL/min) was measured as part of an oral health exam at the UCSF School of Dentistry. Participants were asked to refrain from eating, drinking anything but water, or performing oral hygiene 2 hours prior to oral screening. UWS-FR was collected for all subjects over a period of 5 minutes by a data recorder following a standardized procedure.

In addition, subjects completed 4 surveys to gather information on personal history, medication usage, medical history, and demographical data, and for the visual analog scale (VAS). Study data were collected and managed using REDCap electronic data capture tools hosted at Stanford University (Harris et al. 2019). REDCap (Research Electronic Data Capture) is a secure, web-based software platform designed to support data capture for research studies, providing 1) an intuitive interface for validated data capture; 2) audit trails for tracking data manipulation and export procedures; 3) automated export procedures for seamless data downloads to common statistical packages; and 4) procedures for data integration and interoperability with external sources. The REDCap template for the VAS is included **(Supplementary Data File 1)**.

#### Sample collection sites and protocol

Subjects were asked to refrain from eating, drinking, or performing oral hygiene within 2 hours of sample collection. For each of 3 control and 3 low flow subjects, samples of the buccal and lingual surfaces of index teeth (universal tooth numbers 3, 6 , 8, 9, 11, 14, 19, 22, 24, 25, 27, 30) were collected by a dentist at the UCSF School of Dentistry. Supragingival plaque samples were collected with Epicentre Foam Swabs (Madison, WI, Item #QEC091H). Swabs were applied to either the buccal or lingual tooth aspect in a circular motion with moderate pressure for 5-20 seconds. For subgingival samples, each tooth was sampled independently with two paper points per tooth, one inserted into the mesio-buccal (or mesio-lingual) sulcus and one inserted into the distal-buccal (or distal-lingual) sulcus. Each ISO-45 Paper Point was inserted to the base of the subgingival sulcus or pocket at the mesio-buccal site, until resistance was felt. The paper point was left in place for 20 seconds when it was removed to a sterile collection tube with cotton pliers.

#### DNA extraction and barcoded sequencing of the 16S rRNA gene.

Genomic DNA was extracted from all samples using the MoBio PowerSoil DNA Isolation kit (product #12888–100, Carlsbad, CA) according to the manufacturer's instructions. The V4 region of the 16S rRNA gene was PCR-amplified with barcoded primers, as previously described (DiGiulio et al. 2015; Proctor et al. 2018), pooled in batches of roughly 800 samples per run, and sequenced on the Illumina HiSeq 2500 platform (University of Illinois Roy J. Carver Biotechnology Center, Urbana, IL).

#### Demultiplex and quality filtering

Forward and reverse reads were independently de-multiplexed using the split_libraries_fastq.py command in Qiime 1 with parameters tuned to prevent quality filtering. Sequences were parsed into sample-specific files using the split_sequence_file_on_sample_ids.py command in Qiime before import into R-3.6.1 for quality filtering with the R package dada2 (Callahan et al. 2016); the first 10 nucleotides (5') of each read were trimmed followed by the truncation of forward and reverse reads at lengths of 240 and 160 nucleotides, respectively. Reads were eliminated if the maximum expected error exceeded 2; reads were also truncated at the first instance in the sequence where the quality score was less than 2. Sequences were de-replicated before inference of sequence-specific errors and elimination of problematic reads. Dereplicated and filtered forward sequences were subsequently merged with their paired-end reads before construction of an amplicon sequence variant (ASV) table **(Supplementary Data File 2)**. The removeBimeraDenovo function of dada2 set to the 'consensus' method was used to filter chimeras from each ASV table. Taxonomic assignment was then performed down to the species level where possible using the dada2 implementation of the RDP Naive Bayesian Classifier 3 trained on the RDP database (version 14.0).

#### Decontaminating the ASV table

To identify taxa in the subgingival dataset, we calculated an enrichment score for taxa enriched in controls compared to true samples, as previously described (Proctor et al 2018). A total of 23 technical controls were analyzed in concert with 223 true samples **(Supplementary Data File 3)**. Filtering using this method reduced the data table for the subgingival dataset from 741 to 625 taxa.

#### Impact of clinical variables on community composition.

Constrained correspondence analysis was used to evaluate the extent to which UWS-FR, SWS-FR, and the VAS explained variation in community structure in the microbiome.

#### Estimation of microbial migration

We used neutral community assembly models to estimate microbial migration rates (Munoz, Couteron, and Ramesh 2008) (Sloan et al. 2006). These models assume that the structure of communities is determined by a combination of neutral, or competitively equivalent growth dynamics and random immigration from a shared source "pool" of potential microbial migrants. Neutral community models make few to no assumptions about the physical nature of the source pool (indeed, the microorganisms that make up the source pool may derive from multiple different environments), but rather assume that all habitats are equivalent and that the composition and relative abundances of taxa within the source pool can be approximated by averaging the composition of multiple communities that share the same source pool. Here, we refer to a group of individual, local communities that share a source pool as a "metacommunity".

For our purposes, we defined metacommunities in our dataset as all of the community samples belonging to the same habitat type within an individual (for example, all supragingival, buccal tooth surface samples within a single individual would constitute a metacommunity in our analysis). This was based on our assumption that communities within these groups would both be the most likely to share a source pool, since they inhabit the same mouth, and also be more likely to have similar environmental characteristics, thus increasing the likelihood that differences among them are the result of neutral, rather than selective, dynamics. Thus, we defined four metacommunities for each individual: supragingival buccal, supragingival lingual, subgingival buccal, and subgingival lingual samples, resulting in a total of thirty-two metacommunities across eight subjects.

To estimate migration rates for samples within each of these metacommunities, we used two different methods and compared the results. The first estimates migration rates from measures of community differentiation in a manner analogous to measuring rates of geneflow in populations from measures of population differentiation, such as Fst or Gst (Munoz, Couteron, and Ramesh 2008). The other method estimates migration rates by fitting the data to a neutral community assembly model developed by Sloan et al. (referred to here as the Sloan Neutral Community Model, or SNCM) (Sloan et al. 2006). This model predicts the frequency of occurrence for each taxon, or the proportion of samples in the metacommunity in which each taxon is found, as a function of its abundance in the source pool (approximated by its average abundance across samples in the metacommunity) and a migration rate parameter, which is parameterized by fitting the model to the observed relationship between occurrence frequency and average abundance.

Samples were rarefied to depths ranging between 1000 reads per sample to 50,000 reads per sample at 1,000 read increments in order to determine a depth threshold yielding stable estimates of microbial migration for the Sloan model.

#### Supplementary Tables

**Supplementary Table 1. Demographic composition of patient population.** Race (top), ethnicity (middle) and sex (bottom) of patients broken down by the control and low flow cohorts.

| **Race** | Control Cohort | Low Flow Cohort |
|---|---|---|
| American Indian or Alaskan Native | 4 | 2 |
| Asian | 60 | 9 |
| Black or African American | 7 | 0 |
| Pacific Islander | 1 | 0 |
| White | 44 | 20 |
| Multiracial | 9 | 2 |
| Prefer not to reply | 3 | 3 |

| **Ethnicity** | Control Cohort | Low Flow Cohort |
|---|---|---|
| Hispanic or Latino | 14 | 5 |
| Not Hispanic or Latino | 102 | 26 |
| Prefer not to reply | 3 | 2 |

| **Sex** | Control Cohort | Low Flow Cohort |
|---|---|---|
| Female | 73 | 31 |
| Female to Male Transgender | 0 | 0 |
| Male | 45 | 1 |
| Male to Female Transgender | 0 | 1 |
| Prefer not to answer | 1 | 0 |

Supplementary Table 2. **Analysis of variance on estimated migration rates.** Estimated migration rates were significantly higher in supragingival compared to subgingival habitats (Habitat_Class); however there was no significant difference between buccal and lingual tooth surface habitats (Tooth_Aspect). The effect of cohort (Aim) was also significant. There was a significant interaction between cohort (Aim) and Habitat_Class, indicating the difference between healthy individuals and those with Sjögren's Syndrome was substantial in supragingival habitats but far less pronounced in subgingival habitats.

| | **Df** | **Sum Sq** | **Mean Sq** | **F value** | **p (>F)** |
|---|---|---|---|---|---|
| Habitat_Class | 1 | 0.0073 | 0.0073 | 6.303 | 0.025 |
| Tooth_Aspect | 1 | 0.00E+00 | 0.00E+00 | 0.0278 | 0.8699 |
| Cohort | 1 | 0.0063 | 0.0063 | 5.4572 | 0.0349 |
| Habitat_Class:Tooth_Aspect | 1 | 0.00E+00 | 0.00E+00 | 0.0129 | 0.9111 |
| Habitat_Class:Cohort | 1 | 0.0052 | 0.0052 | 4.4883 | 0.0525 |
| Tooth_Aspect:Cohort | 1 | 0.00E+00 | 0.00E+00 | 0.008 | 0.9299 |
| Habitat_Class:Tooth_Aspect:Cohort | 1 | 0.00E+00 | 0.00E+00 | 0.0032 | 0.9556 |
| Residuals | 14 | 0.0162 | 0.0012 | | |

### Supplementary Figures

#### Supplementary Figure 1

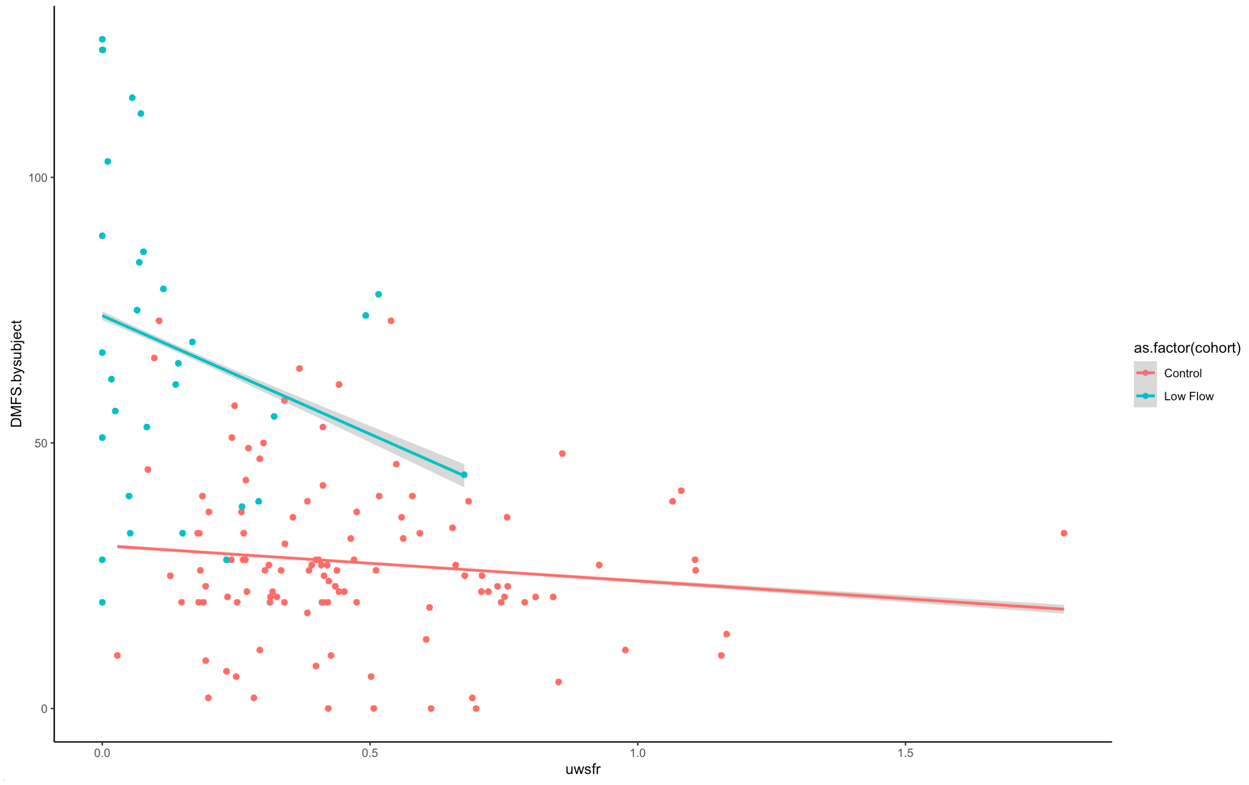

**Supplementary Figure 1. Scatterplot of DMFS as a function of UWS-FR.** Linear models were fit for each cohort. As flow rate decreased, the number of DMFS increased at a greater rate in the low flow cohort (blue) compared to controls (orange).

#### Supplementary Figure 2

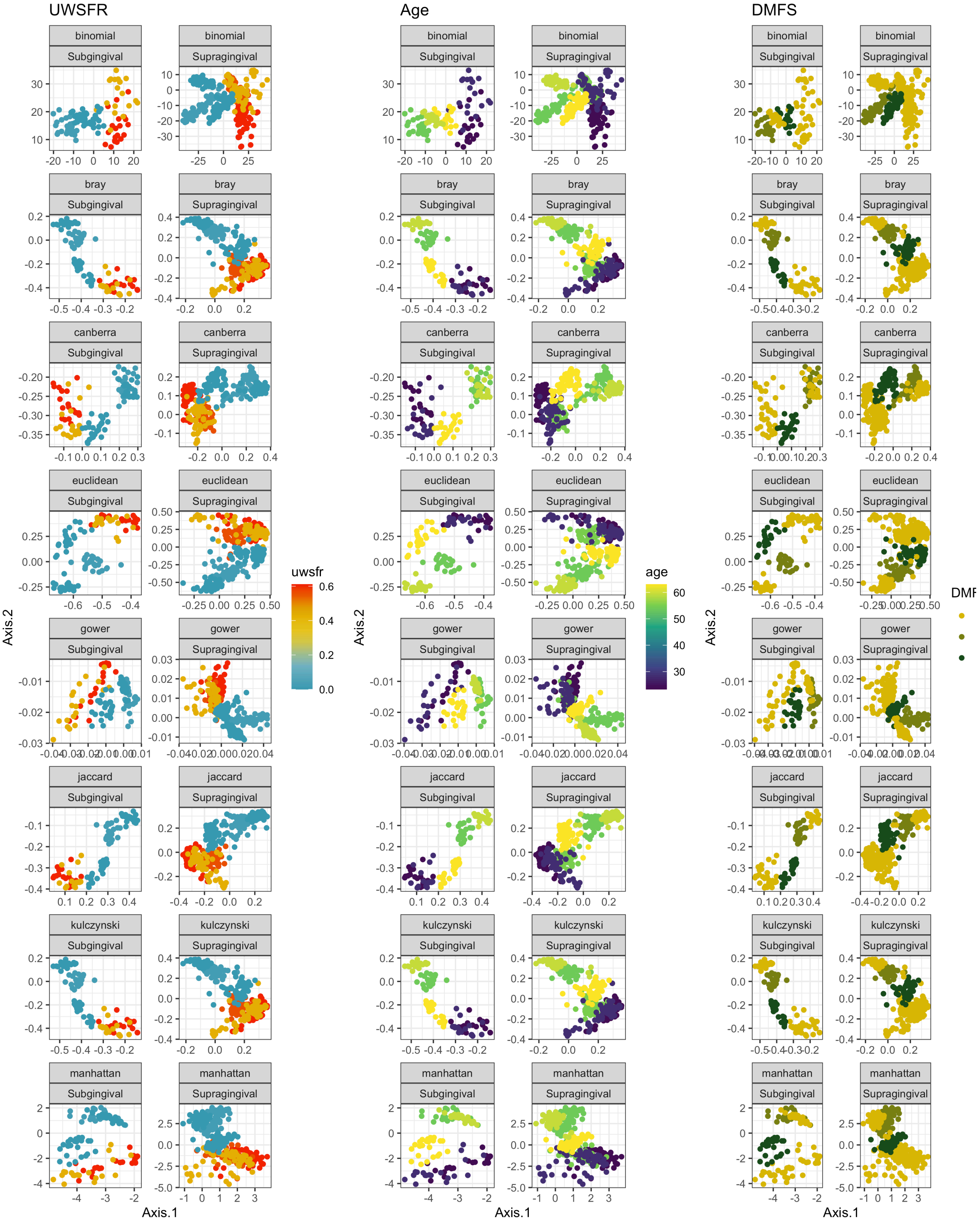

**Supplementary Figure 2. Patterns observed in Figure 3 can be seen using a variety of different ecological distance metrics.** Principal coordinates analysis on various dissimilarity metrics (bray, Canberra, Euclidean, Manhattan, kulzynski, gower) using the combined subgingival and supragingival dataset. Each independent PCoA on each distance metric is displayed as a facet wrap with subgingival and supragingival samples in left and right panels, respectively. Left panels represent samples shaded by UWS-FR, middle panels are shaded by participant age, and right panels are shaded by the average DMFS per tooth.

#### Supplementary Figure 3

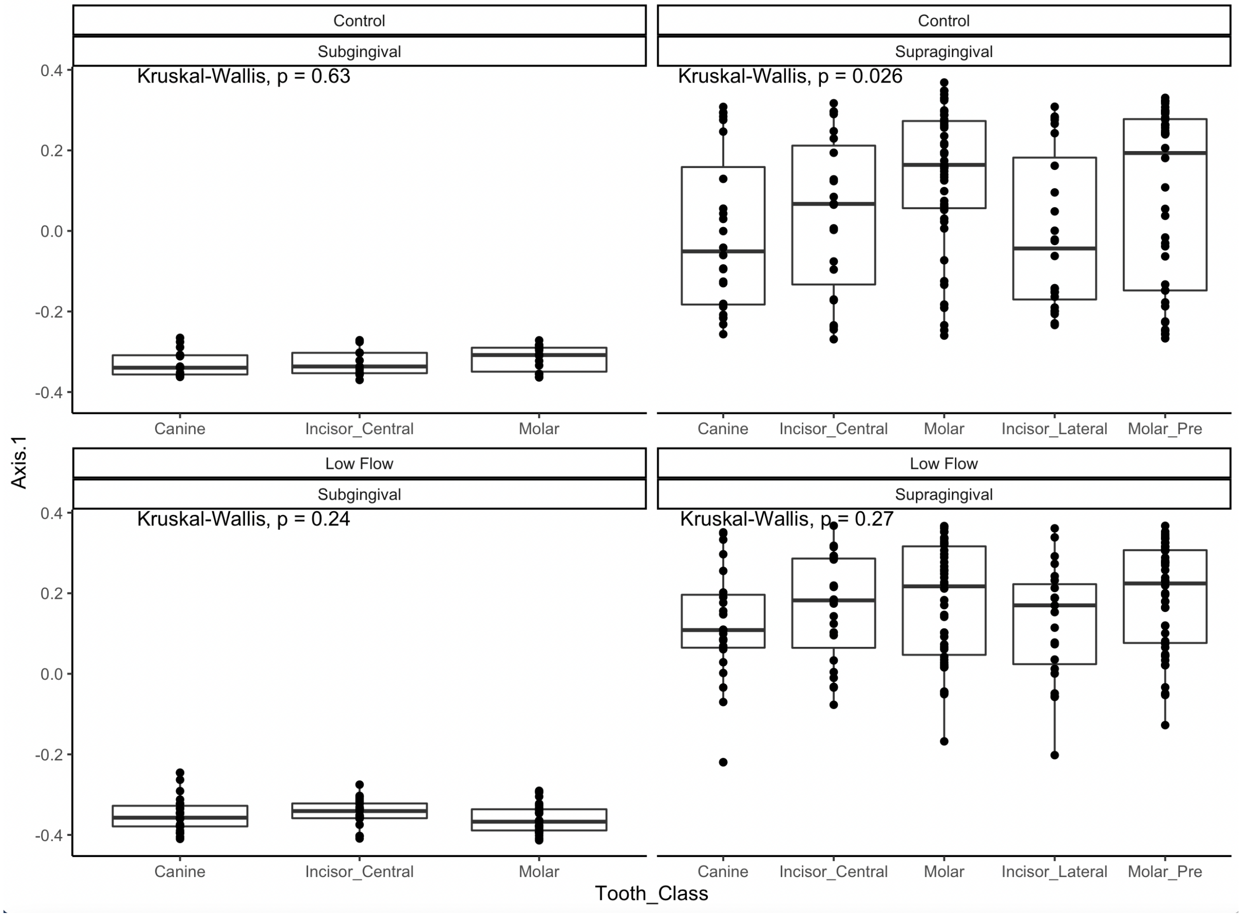

**Supplementary Figure 3. Axis 1 scores plotted as a function of tooth class.** Supragingival communities differed significantly in the control cohort, compared to the low flow subjects. Site-to-site variability was not observed for subgingival sites in either cohort.

#### Supplementary Figure 4

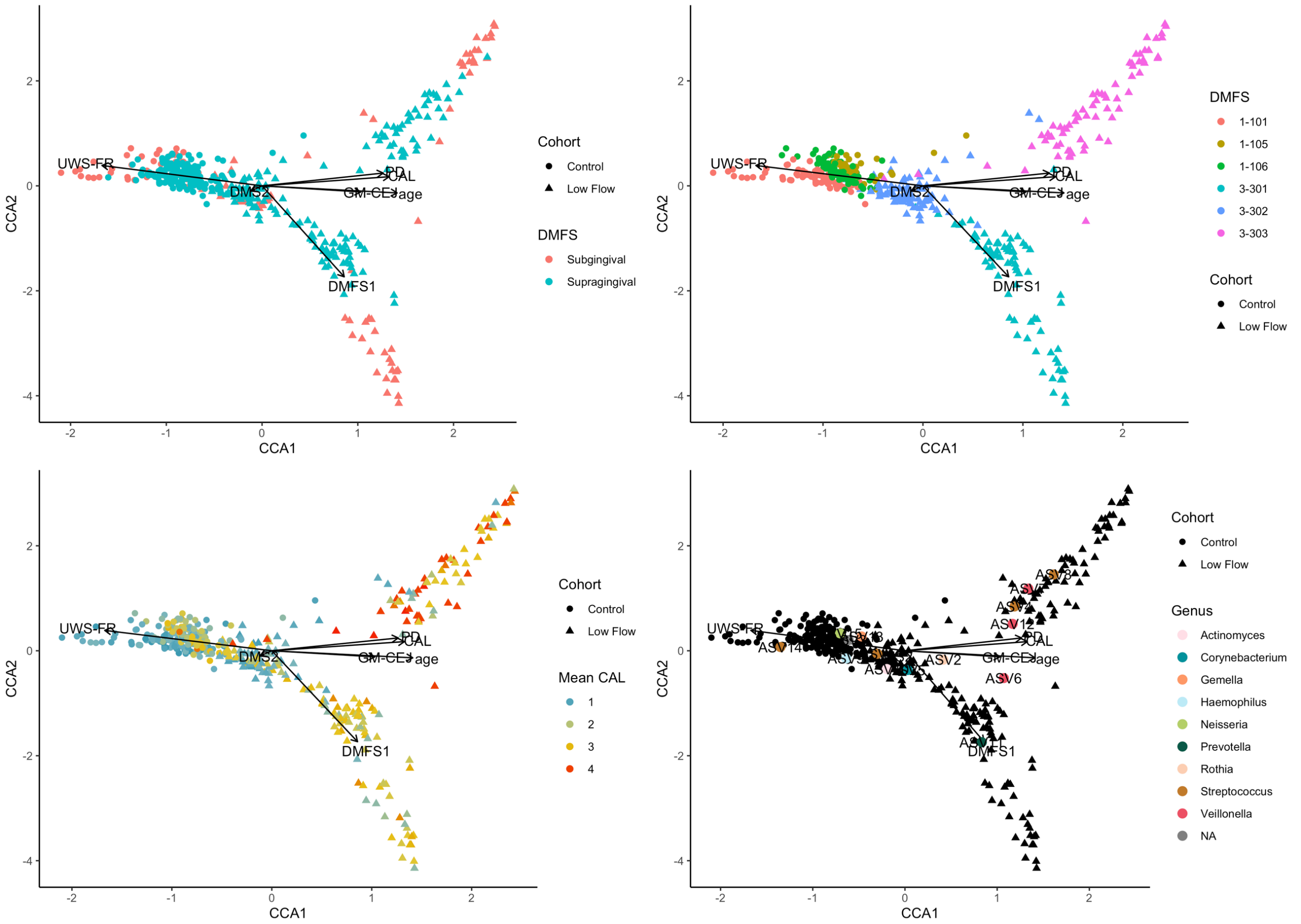

**Supplementary Figure 4. Constrained correspondence analysis of microbial data using clinical variates as predictors.** Canonical correlation shows that age, pd, and dmfs are correlated.

#### Supplementary Figure 5

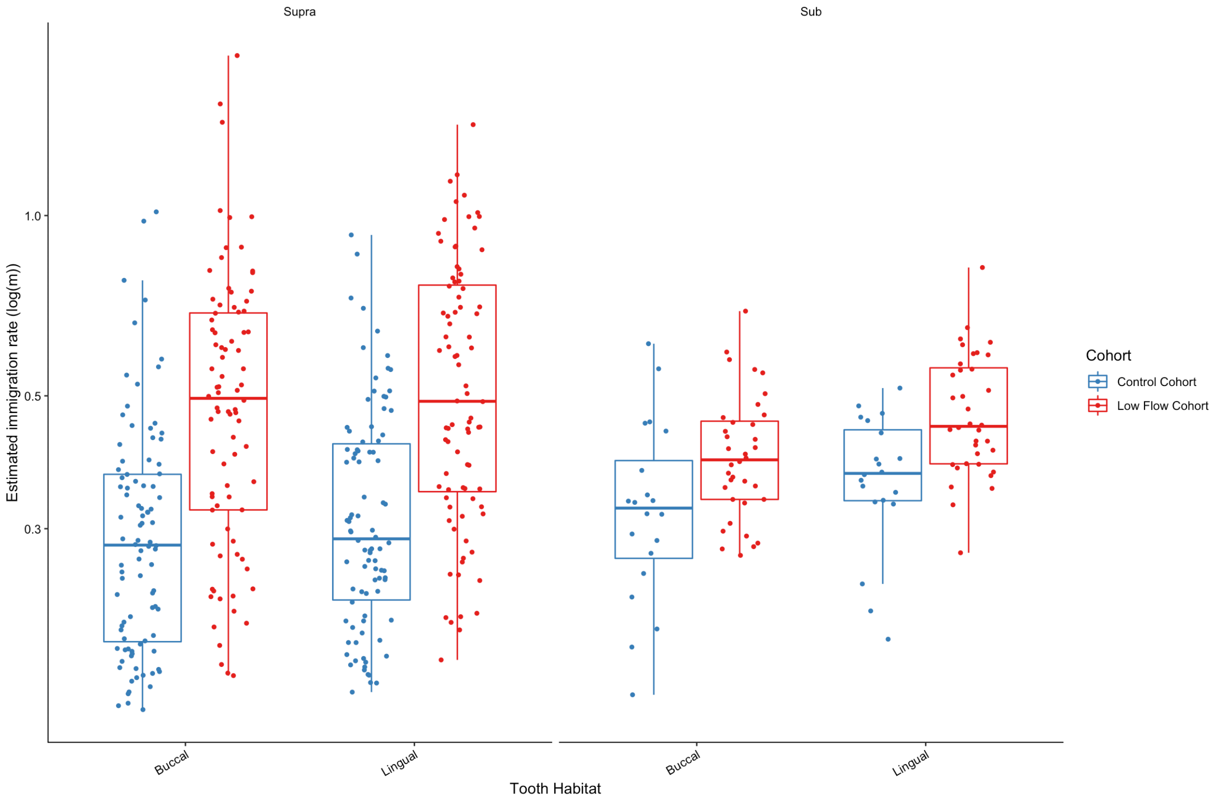

**Supplementary Figure 5. Estimated migration rates (Gst) for each tooth habitat and tooth aspect.** Colors correspond to cohort (control cohort; low flow cohort). In these boxplots, outer edges of boxes encompass the first and third quartiles. The horizontal line bisecting the center of each box demarcates the median. Each point represents an individual sample/community within each of the defined metacommunities.

#### Supplementary Figure 6

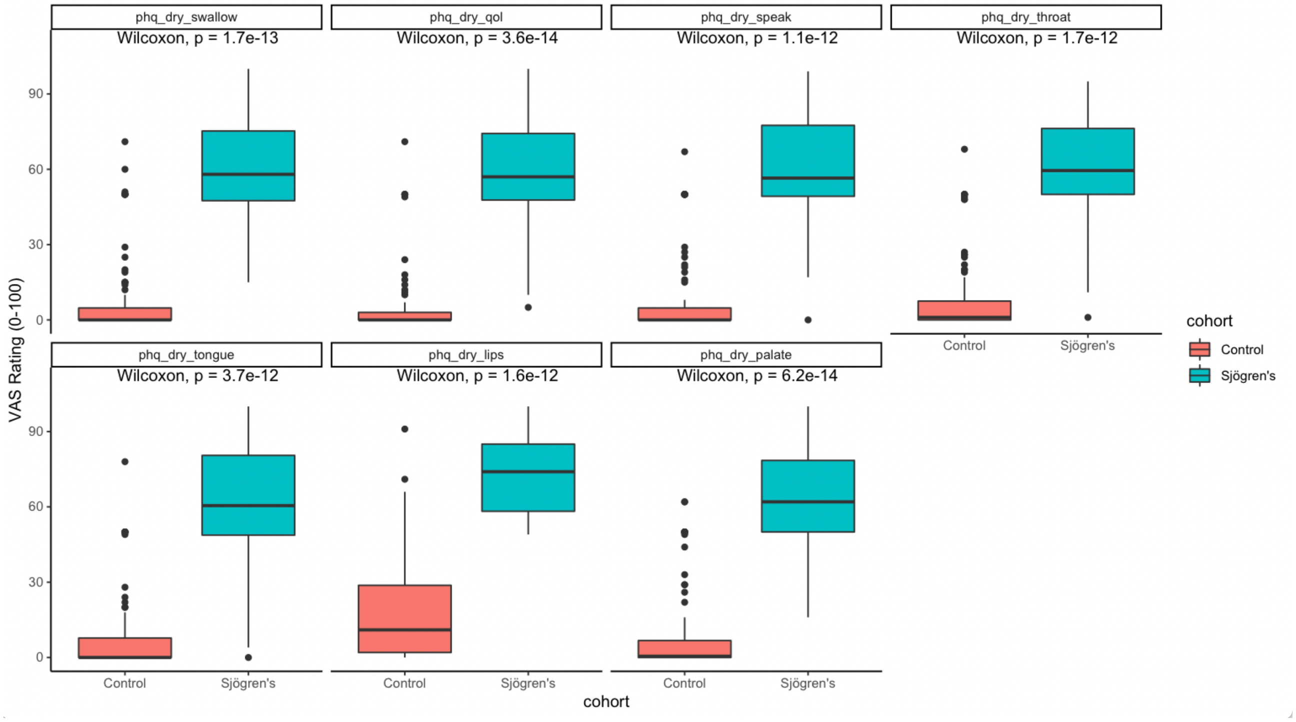

**Supplementary Figure 6. Boxplots of VAS reveal significant differences in responses between groups**. Each question on the Visual Analog Scale querying the symptomatic complaints of dry mouth is plotted as an individual panel. The midline of each boxplot represents the median while the outer edges encompass the first and third quartiles. Points on the graph indicate outliers. Colors correspond to patient cohort (control, low flow). Across all questions, respondents in the low flow cohort rated their impairment and subjective complaints of dry mouth as more severe than the patients in the control cohort (Wilcoxon rank sum tests, $p < 0.05$).

#### Supplementary Figure 7

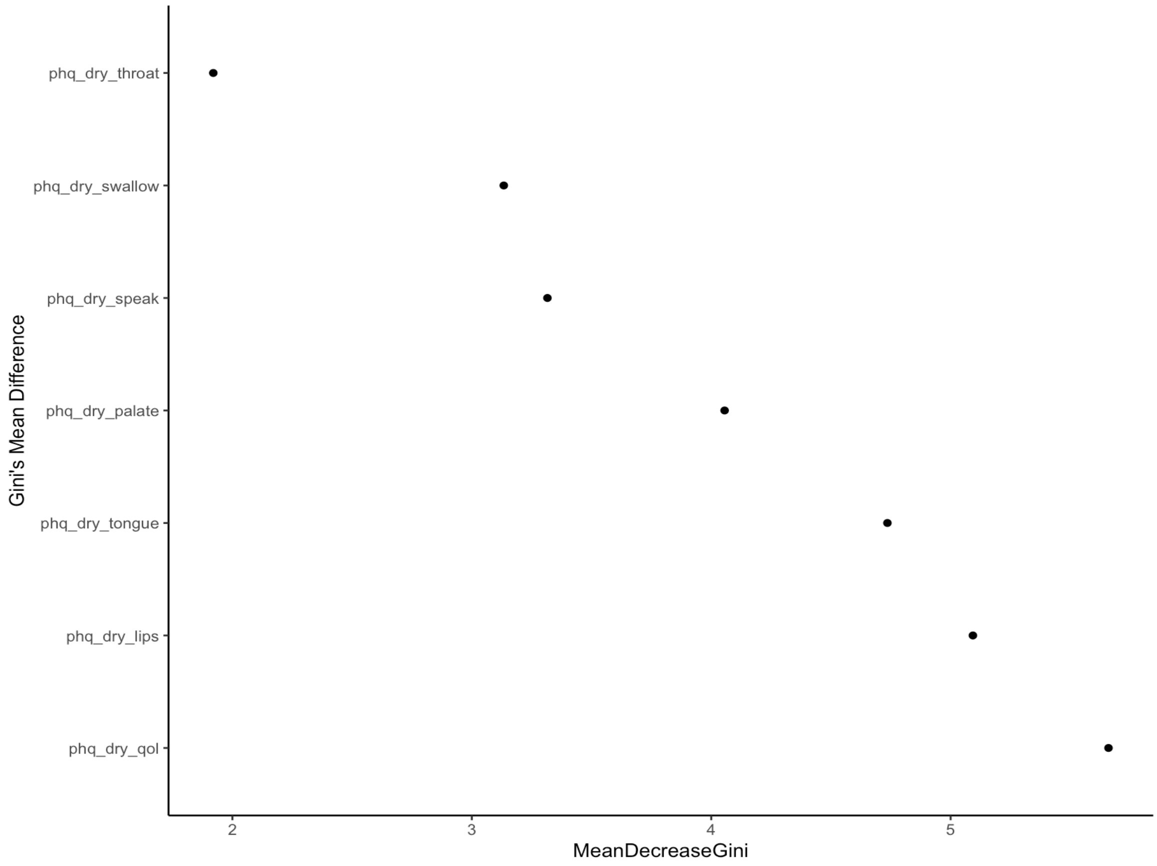

**Supplementary Figure 7. Random forest analysis identifies dryness of lips and impacts on quality of life as the most discriminant features.** Random forest analysis was used to identify the most discriminant features included in the Visual Analog Scale. The out-of-box error rate for the random forest model was 8.46%. The most discriminant features were the impact of dry mouth on the overall quality of life (phq_dry_qol), and the dryness of the lips (phq_dry_lips) consistent with the classification tree.
